# Ticagrelor vs Clopidogrel: the Impact of Platelet Inhibition on Cerebrovascular Microembolic Events during TAVR

**DOI:** 10.1101/2020.11.19.20234377

**Authors:** Michael A. Vavuranakis, Charalampos Kalantzis, Vassilis Voudris, Elias Kosmas, Konstantinos Kalogeras, Efstratios Katsianos, Evaggelos Oikonomou, Gerasimos Siasos, Konstantinos Aznaouridis, Konstantinos Toutouzas, Myrsini Stasinopoulou, Argyro Tountopoulou, Evangelia Bei, Carmen M. Moldovan, Dimitrios Vrachatis, Ioannis Iakovou, Theodore G. Papaioannou, Dimitrios Tousoulis, Thorsten M. Leucker, Manolis Vavuranakis

## Abstract

**Objectives:** To evaluate the effects of ticagrelor versus clopidogrel and of platelet inhibition on the number of cerebrovascular microembolic events, in patients undergoing transcatheter aortic valve replacement (TAVR).

**Background:** The impact of the antiplatelet regimen and the extent of associated platelet inhibition on cerebrovascular microembolic events during TAVR are unknown.

**Methods:** Patients scheduled for TAVR were randomized prior to the procedure to either aspirin and ticagrelor or to aspirin and clopidogrel. Platelet inhibition was expressed in P2Y12 Reaction Units (PRU) and percentage of inhibition. High intensity transient signals (HITS) were assessed with transcranial Doppler (TCD). Safety outcomes were recorded according to the VARC-2 definitions.

**Results:** Among 90 patients randomized, six had inadequate TCD signal. The total number of procedural HITS was lower in the ticagrelor group (416.5 [324.8, 484.2]) (42 patients) than in the clopidogrel group (723.5 [471.5, 875.0]) (42 patients), p< 0.001. After adjusting for the duration of the procedure, diabetes, extra-cardiac arteriopathy, BMI, and aortic valve calcium content, patients on ticagrelor had on average 255.9 (95% CI: [-335.4, -176.4]) fewer total procedural HITS, than did patients on clopidogrel. Platelet inhibition was greater in those randomized to ticagrelor 26 [10, 74.5] PRU than in those randomized to clopidogrel 207.5 [120-236.2] PRU, p<0.001 and correlated significantly with procedural HITS (r=0.5, p<0.05). This protective effect was not associated with an increase in complications.

**Conclusions:** Ticagrelor resulted in fewer procedural HITS, compared to clopidogrel, in patients undergoing TAVR, while achieving greater platelet inhibition, without increasing the risk for complications.

**Clinical Trial:** **(ClinicalTrials.gov Identifier: NCT02989558)**

**CONDENSED ABSTRACT:** We conducted a two-center, prospective, open label, randomized, controlled clinical trial to compare the efficacy of ticagrelor vs clopidogrel in preventing cerebrovascular embolic events as assessed by transcranial Doppler during TAVR.

The total number of procedural HITS was lower in the ticagrelor group (416.5 [324.8, 484.2]) than in the clopidogrel group (723.5 [471.5, 875.0]), p< 0.001. Patients on ticagrelor had on average 255.9 (95% CI: [-335.4, -176.4]) fewer total procedural HITS than those on clopidogrel. This protective effect was not associated with an increase in complications.

## INTRODUCTION

Transcatheter Aortic Valve Replacement (TAVR) provides a therapeutic alternative to surgical valve replacement for patients with symptomatic severe aortic stenosis (AS) (1). Cerebrovascular events are among the most clinically significant complications of TAVR (2). Magnetic Resonance Imaging (MRI) studies have documented the development of new cerebral infarct lesions in up to 70% of patients undergoing TAVR, most of which are not recognized at the time of the procedure (3). Transcranial Doppler (TCD) is a valuable tool for the detection of clinically “silent” embolic cerebrovascular events during TAVR (4).

The source of cerebral embolic infarcts during TAVR is considered multifactorial (5, 6). Inadequate platelet inhibition during or after the procedure, may increase the risk of thrombus formation both on the mechanically eroded aortic atherosclerotic plaques and/or the bioprosthetic valve frame and leaflet tissue (6) and platelet inhibition during the peri- and post TAVR periods may decrease the incidence of these events. The relative risks and benefits of anti-platelet regimens in TAVR patients are not well characterized (7, 8). Ticagrelor results in more potent platelet inhibition than does clopidogrel (9). In addition, it reduces cerebral ischemic events in patients with acute coronary syndromes (ACS) without significantly increasing major bleeding complications (10). A recent study reported that the ticagrelor and aspirin regimen was able to achieve sufficient platelet reactivity as defined by PRU <208 in the 70% of the studied post-TAVR population who had failed to do so with clopidogrel and aspirin (11). However, the relative impacts of the extent of platelet inhibition achieved by ticagrelor and clopidogrel on the incidence of TCD-defined microemboli and of bleeding in the TAVR patient population have not been studied.

The present study evaluated whether platelet inhibition with a more potent antiplatelet agent would provide better cerebral protection from microembolizations in the cerebral circulation during TAVR. We hypothesized that treatment with aspirin and ticagrelor in patients undergoing TAVR results in fewer high-intensity transient signals (HITS) as assessed by TCD of the middle cerebral arteries than does treatment with aspirin and clopidogrel.

## METHODS

### Study design and patient population

The PTOLEMAIOS trial (ClinicalTrials Identifier: NCT02989558) was a two-center, prospective, open label, randomized, controlled clinical trial. Consecutive patients with symptomatic severe native aortic valve stenosis, who were deemed at high risk for surgical aortic valve replacement (SAVR) (logistic EuroSCORE ≥ 18) or inoperable, were included in the trial. We excluded patients with a history of atrial fibrillation, any coagulopathy disorder (acquired or congenital), patients receiving anticoagulation or who had received antiplatelet therapy other than aspirin within 7 days before randomization and patients with significant carotid artery disease (>50% on ultrasonography). The inclusion and exclusion criteria are presented in detail in the **Supplementary Appendix**.

The study was conducted in accordance with the ethical principles for medical research of the Declaration of Helsinki, International Conference on Harmonization (ICH) /Good Clinical Practice (GCP), the European Union Clinical Trials Directive, and Greek legislation. The study protocol was approved by the hospitals’ Ethics Committee and Institutional Review Board, the National Ethics Committee (NEC), and the National Organization of Medicines (NOM). Safety updates were provided according to local requirements, including SUSARs (Suspected Unexpected Serious Adverse Reactions).

The study protocol was divided into 5 stages:

Baseline (days -10 to -1 from TAVR date)

A complete TAVR evaluation screening was performed in patients meeting eligibility criteria (see **Appendix B-Flowchart of Study Procedures):**

All patients received aspirin 80 mg daily for 7 days prior to the TAVR procedure, followed by the same dose for the next 90 days. In addition, patients were randomized to clopidogrel at a loading dose of 300 mg 1 day prior to the procedure, and 75 mg daily for the following 90 days (Group 1) or to ticagrelor 90 mg twice daily 1 day prior to the procedure and for the following 90 days (Group 2).

### TAVR Procedure (Day 0)

Platelet inhibition was assessed with the VerifyNow point of care (Accumetrics, San Diego CA, USA) on the day of the TAVR procedure. For all patients, TAVR was performed via femoral access with a self-expandable valve. Unfractionated heparin was used for anticoagulation with procedural ACT ≥ 300. An arterial closure device (Perclose Proglide, Abbot, Chicago IL, USA) was used for hemostasis. When deemed necessary predilation of the native aortic valve was performed.

TCD recordings were obtained in all patients for 10 minutes prior to and throughout the procedure. The procedure was separated into 4 phases **(see Supplementary Appendix)**. Quality of TCD signals was verified by a device operator blinded to the antiplatelet regimen. Adverse events (AE) and serious adverse events (SAE) were recorded.

### Discharge (Days 4 to 7)

Prior to discharge, participants underwent a physical examination, neurological assessment, evaluation of platelet inhibition, and a 30-minute TCD recording AEs, SAEs and bleeding events were recorded.

### End of Treatment (Day 90)

Subjects were evaluated for compliance, AEs, SAEs and bleeding events and a physical examination and neurological assessment were performed.

### End of Study

A scheduled telephone or, when possible, an in-person follow-up was performed 120 (± 5) days after TAVR. Participants were evaluated regarding compliance, AEs, SAEs and bleeding events. If the subject had taken the investigational products for more than 80% of the days, they were considered compliant.

### Study procedures

#### Transcranial Doppler (TCD)

The Digi-Lite™ (Rimed Inc, Long Island NY, USA) TCD device with two bilateral 2 MHz probes was used to sonicate the middle cerebral arteries at a depth of 48 mm-56 mm with a sample volume of 8 mm-12 mm.

TCD was performed for 10 minutes prior to TAVR, throughout the TAVR procedure, and for 30 minutes on the discharge day by two operators blinded to the patient’s randomization group. For consistency, the setup, recordings, and analysis for the baseline and follow-up TCD studies for each participant were performed by the same operator.

#### HITS detection

HITS detection was performed automatically using multi-depth embolic detection with artifact rejection. The lowest threshold for discriminating microembolic signals from the background noise (3 dB) was selected (12). The total number of HITS during the duration of the TAVR procedure were analyzed.

#### Neurologic Assessment

A neurologic assessment by a blinded neurologist was performed at baseline, discharge, and at 90 days after TAVR. Assessment included motor, sensory, cranial nerve, and cerebellar function tests and a Mini-Mental Status examination (13).

#### Assessment of residual platelet reactivity

Residual platelet reactivity was evaluated for all study subjects using the VerifyNow P2Y12 assay (Accumetrics Inc, USA) before the procedure and at discharge. P2Y12 receptor blockade was measured in P2Y12 reaction units (PRU) and percentage of inhibition. In addition, the base PRU, a P2Y12-independent measurement of platelet function based on the rate and extent of platelet aggregation from the thrombin receptors, specifically the PAR-1 and PAR-4 receptors, was measured. The study staff analyzing the samples was blinded to the subjects’ treatment group.

### Study Outcomes

#### Primary Outcome

The primary outcome was the difference in the number of HITS detected by TCD throughout the TAVR procedure between the subjects receiving ticagrelor and those receiving clopidogrel.

#### Secondary Outcomes

The study’s secondary outcomes included:

i. The differences in the incidence of HITS between the two treatment groups for each phase of the TAVR, and at discharge.
ii. to compare residual platelet reactivity among the two groups, at the day of the procedure and at hospital discharge.
iii. to access the relationship between the extent of platelet inhibition and the number of HITS.

### Safety Outcomes

The safety outcomes were the AEs, SAEs, and bleeding events, as defined by VARC-2 criteria, perioperatively and at 90 days post TAVR in the two groups. Definitions of AEs and SAEs can be found in the (**Supplementary Appendix**). Bleeding events were recorded according to VARC-2 definitions (14) up to 90 days post TAVR. Cerebrovascular events were recorded according to VARC-2 definitions up to 90 days post TAVR and were confirmed by a board-certified neurologist. Minor stroke was defined as a Modified Rankin score < 2 and a major stroke if the Modified Rankin score was ≥2 (15).

### Statistical analysis

Continuous variables are presented as mean ± standard deviation or median [Q1, Q3] and compared using the Welch two sample t-test or the Wilcoxon rank sum test, as appropriate. The Wilcoxon Signed Rank Test was used for within group comparisons. Categorical variables are presented as counts (percentages) and compared using the Fisher’s exact test.

The Intent to Treat Population (ITT) included all randomized subjects who received study medication. Safety outcomes were analyzed using the ITT population.

Multiple linear regression analysis was used to quantify the effect of ticagrelor compared to clopidogrel on the total number of procedural HITS. Among age, gender, CHA_2_DS_2_VASc score, antiplatelet therapy, and duration of the procedure, only the antiplatelet therapy and procedure duration had a significant correlation to HITS and were added to the final model. Potential confounders (diabetes, hypertension, extracardiac arteriopathy, carotid atherosclerotic disease/peripheral vascular disease, aortic valve calcium content, pre-implantation aortic balloon valvuloplasty) as well as baseline BMI (which was significantly different between the two groups at baseline) were added to the final model to obtain an adjusted effect size. Initial PRU was not included due to collinearity with antiplatelet therapy. Linear regression models with and without additional covariates revealed no confounding.

Two additional models, using the same covariates, were used to assess the effect of PRU and percent of platelet inhibition on HITS. Finally, to assess the effect of antiplatelet regimen on PRU, we fit a linear regression model with PRU as the outcome variable including covariates known to affect PRU such as sex, platelet count, hematocrit, and BMI (16).

The Bland-Altman method was used to depict intra-observer variability and inter-observer difference. All statistical tests were two-sided with a significance level set at 0.05. Statistical analysis was performed using the RStudio Team (2019), Boston, MA.

### Sample size and power calculations

Size of study population was calculated utilizing the formula by Pocock SJ (17) (**Supplementary Appendix**).

## RESULTS

### Demographic and Clinical Characteristics (Table 1)

During the study period, 150 consecutive patients with severe symptomatic AS were evaluated for participation in the study. Sixty patients were excluded based on exclusion criteria (see Appendix). Ninety patients were randomized to either ticagrelor (45 participants) or to clopidogrel (45 participants). Six patients were excluded from the final analysis because of an inadequate TCD acoustic window or a poor signal **(Figure 1)**.

**Table 1.** Baseline Demographic and Clinical characteristics of the trial population.

|  | Intention to Treat Population |  |  | P value | As Treated Population |  |  | P value |
| --- | --- | --- | --- | --- | --- | --- | --- | --- |
|  | Total population (N=90) | ASA & Clopidogrel (N=45) | ASA & Ticagrelor (N=45) |  | Total population (N= 84) | ASA & Clopidogrel (N=42) | ASA & Ticagrelor (N=42) |  |
| Age (years) | 82 ± 8 | 82.3 ± 9.6 | 81.6 ± 6.1 | 0.09 | 82.06 ± 8 | 82.5 ± 9.5 | 81.6 ± 6.2 | 0.09 |
| Male gender, n (%) | 56 (62.2) | 28 (62.2) | 28 (62.2) | 1 | 53 (63.1) | 26 (61.9) | 27 (64.3) | 1 |
| BMI (kg/m<sup>2</sup>) | 26.7 ± 4 | 25.8 ± 3.3 | 27.7 ± 4.3 | 0.02 | 26.7 ± 4 | 25.7 ± 3.4 | 27.7 ± 4.4 | 0.02 |
| BSA (m<sup>2</sup>) | 1.9 ± 0.4 | 1.9 ± 0.3 | 1.9 ± 0.4 | 0.31 | 1.9 ± 0.4 | 1.8 ± 0.3 | 1.9 ± 0.4 | 0.32 |
| COPD, n (%) | 33 (36.7) | 16 (35.6) | 17 (37.8) | 1 | 32 (38) | 16 (38.1) | 16 (38.1) | 1 |
| Previous PCI, n (%) | 27 (30) | 14 (31.1) | 13 (28.9) | 1 | 27 (32.1) | 14 (33.3) | 13 (31) | 1 |
| Previous cardiac surgery, n (%) | 20 (22.2) | 10 (22.2) | 10 (22.2) | 1 | 17 (20.2) | 8 (19) | 9 (21.4) | 1 |
| Creatinine < 2mg/dl, n (%) | 81 (90) | 39 (86.7) | 42 (93.3) | 0.48 | 75 (89.3) | 36 (85.7) | 39 (92.9) | 0.48 |
| Extracardiac arteriopathy, n (%) | 28 (31.1) | 11 (24.4) | 17 (37.8) | 0.25 | 26 (31) | 10 (23.8) | 16 (38.1) | 0.24 |
| Preexisting Neurological dysfunction, n (%) | 7 (7.8) | 4 (8.9) | 3 (6.7) | 1 | 7 (8.3) | 4 (9.5) | 3 (7.1) | 1 |
| Coronary artery Disease, n (%) | 51 (56.7) | 28 (62.2) | 23 (51.1) | 0.4 | 48 (57.1) | 26 (62) | 22 (52.4) | 0.51 |
| History of Myocardial Infarction, n (%) | 1 (1.1) | 0 (0) | 1 (2.2) | 1 | 1 (1.2) | 0 (0) | 1(2.4) | 1 |
| Smoking, n (%) | 30 (33.3) | 14 (31.1) | 16 (35.5) | 0.82 | 28 (33.3) | 13 (30.9) | 15 (35.7) | 0.82 |
| Hypertension, n (%) | 76 (84.4) | 39 (86.7) | 37 (82.2) | 0.77 | 70 (83.3) | 36 (85.7) | 34 (81) | 0.77 |
| Hyperlipidemia, n (%) | 70 (77.8) | 31 (68.9) | 39 (86.7) | 0.07 | 66 (78.6) | 30 (71.4) | 36 (85.7) | 0.18 |
| Diabetes, n (%) | 25 (27.8) | 12 (26.7) | 13 (28.9) | 1 | 23 (27.4) | 11 (26.2) | 12 (28.6) | 1 |
| EuroSCORE II | 5.69 ± 3.99 | 6.20 ± 3.99 | 5.18 ± 3.97 | 0.23 | 5.7 ± 4.1 | 6.3 ± 4.1 | 5.2 ± 4.1 | 0.2 |
| STS score | 5.28± 3.64 | 5.53 ± 2.83 | 5.03 ± 4.32 | 0.52 | 5.3 ± 3.7 | 5.7 ± 2.9 | 4.9 ± 4.3 | 0.36 |
| Previous Permanent Pacemaker, n (%) | 12 (13.3) | 6 (13.3) | 6 (13.3) | 1 | 12 (14.3) | 6 (14.3) | 6 (14.3) | 1 |
| Previous Valvuloplasty, n (%) | 10 (11.1) | 4 (8.9) | 6 (13.3) | 0.74 | 8 (9.5) | 3 (7.1) | 5 (11.9) | 0.71 |
| HAS-BLED score | 2.58 ± 0.72 | 2.60 ± 0.69 | 2.56 ± 0.76 | 0.77 | 2.58 ± 0.73 | 2.6 ± 0.7 | 2.57 ± 0.77 | 0.88 |
| CHA<sub>2</sub>DS<sub>2</sub>VASc score | 4.57 ± 1.15 | 4.58 ± 1.08 | 4.56 ± 1.24 | 0.93 | 4.58 ± 1.17 | 4.62 ± 1.08 | 4.55 ± 1.27 | 0.78 |

Continuous variables are reported as mean±SD.
ASA= acetylsalicylic acid; SD = standard deviation; BMI = body mass index; BSA= body surface area; COPD= chronic obstructive pulmonary disease; PCI= percutaneous coronary intervention

**Figure 1.**
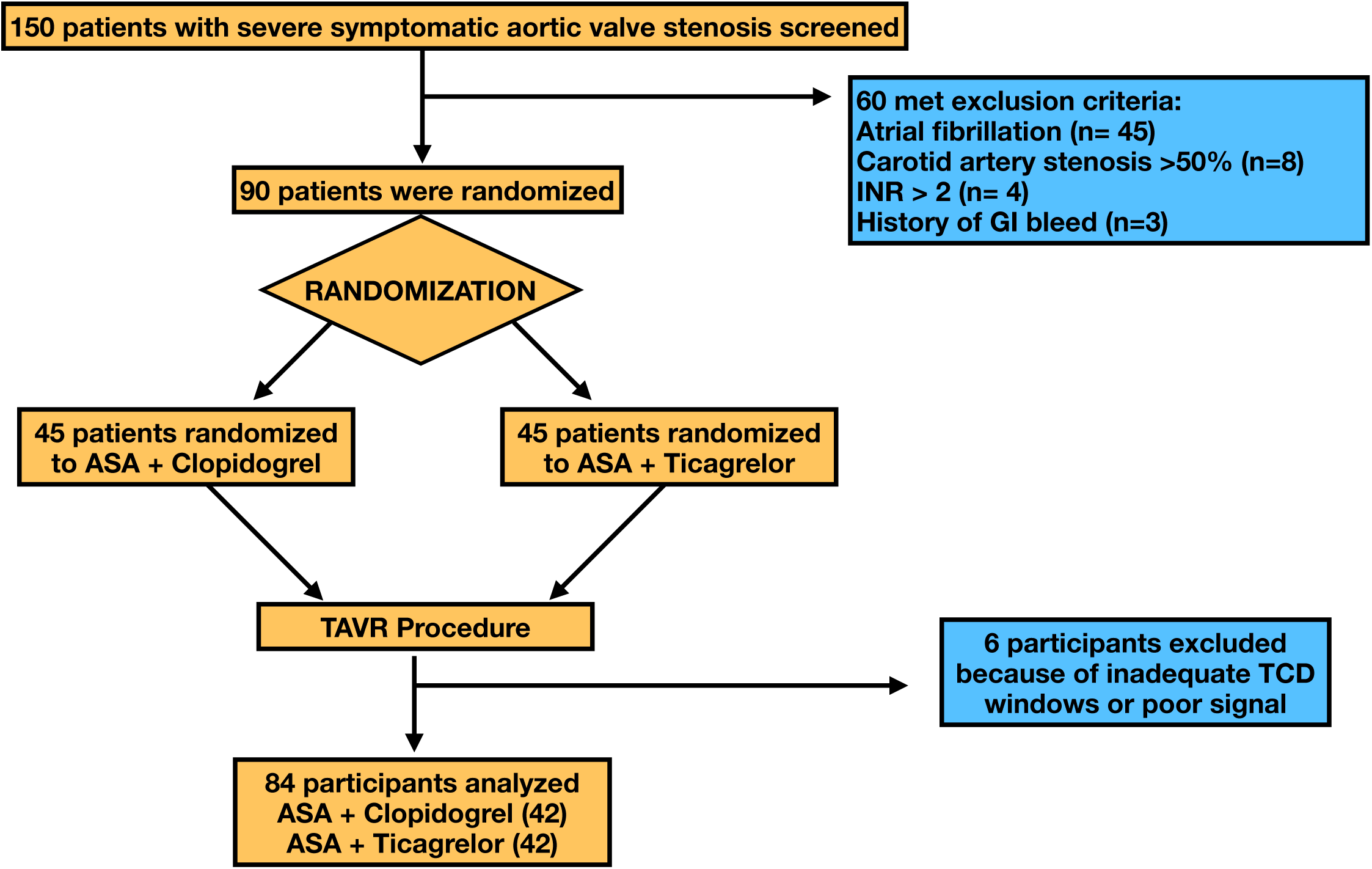
PTOLEMAIOS Study Flow Chart. ASA= acetylsalicylic acid; TAVR= Transcatheter Aortic Valve Replacement; TCD= Transcranial Doppler Study.

In the intention to treat population, there were no statistically significant differences in the demographic and clinical characteristics with the exception of body mass index (BMI), which was higher in the ticagrelor group. In 84 patients [aspirin and clopidogrel (42 patients), aspirin and ticagrelor (42 patients)] good quality TCD recordings were obtained during the procedure and were analyzed and, in these patients as well, the baseline demographic and clinical characteristics were evenly distributed, except for BMI which was higher in the ticagrelor group. Therefore, BMI was adjusted for in the regression analysis.

### Procedural data

All 84 patients underwent a successful TAVR procedure with a self-expandable valve (Evolut R, Medtronic, Dublin, Ireland). Procedural characteristics were evenly distributed with no significant group differences **(Table 2)**.

**Table 2.** Procedural Characteristics.

|  | Total<br>Population<br>(n= 84 ) | ASA + Clopidogrel<br>(n= 42 ) | ASA+ Ticagrelor<br>(n= 42 ) | p value |
| --- | --- | --- | --- | --- |
| Procedure Duration (minutes) | 87.6 ± 21.2 | 88 ± 21.6 | 87.2 ± 21.1 | 0.9 |
| Phase 1 | 41.4 ± 14.2 | 41.9 ± 12.3 | 40.9 ± 16.1 | 0.75 |
| Phase 2 | 6.4 ± 5.3 | 6.6 ± 4.8 | 6 ± 6.1 | 0.8 |
| Phase 3 | 14.8 ± 6.6 | 14.6 ± 6.5 | 14.9 ± 6.9 | 0.8 |
| Phase 4 | 29.9 ± 14.2 | 29.7 ± 15.7 | 30.1 ± 12.8 | 0.9 |
| Number of total Angiographic Injections | 10.31 ± 2.9 | 10.37 ± 2.3 | 10.24 ± 3.5 | 0.9 |
| General Anesthesia, n (%) | 34 (40.5) | 13 (31) | 22 (50) | 0.08 |
| Balloon Pre-dilatation (phase 2), n (%) | 20 (23.8) | 11 (26.2) | 9 (21.4) | 0.16 |

Continuous variables are reported as mean± SD.

### Primary outcome

The total number of procedural HITS was significantly lower in the ticagrelor group than in the clopidogrel group (416.5 [324.8, 484.2] vs 723.5 [471.5, 875.0] respectively, p <0.001) **(Table 3) (Figure 2-Central Illustration)**.

**Table 3.** Number of High Intensity Transient Signals during the TAVR Procedure

|  | Total Population<br>(n= 84) | ASA + Clopidogrel<br>(n= 42) | ASA + Ticagrelor<br>(n= 42) | p value |
| --- | --- | --- | --- | --- |
| Phase 1 | 208 [134, 331.2] | 289 [161.5, 412.8] | 170 [99.75, 222.50] | <0.001* |
| Phase 2 | 36 [27, 57] | 47 [36, 75] | 24 [22, 33] | 0.012* |
| Phase 3 | 147.50 [100.5, 211.8] | 184 [111.5, 266.8] | 142 [90.75, 168.50] | 0.024* |
| Phase 4 | 103.50 [53.75, 158.25] | 130 [57, 232.8] | 89 [43.25, 128.50] | 0.011* |
| All Phases | 479 [380.2, 720.2] | 723.50 [471.5, 875.0] | 416.50 [324.8; 484.2] | <0.001* |

High intensity transient signals reported as median [quartile 1, quartile 3].

**Figure 2.**
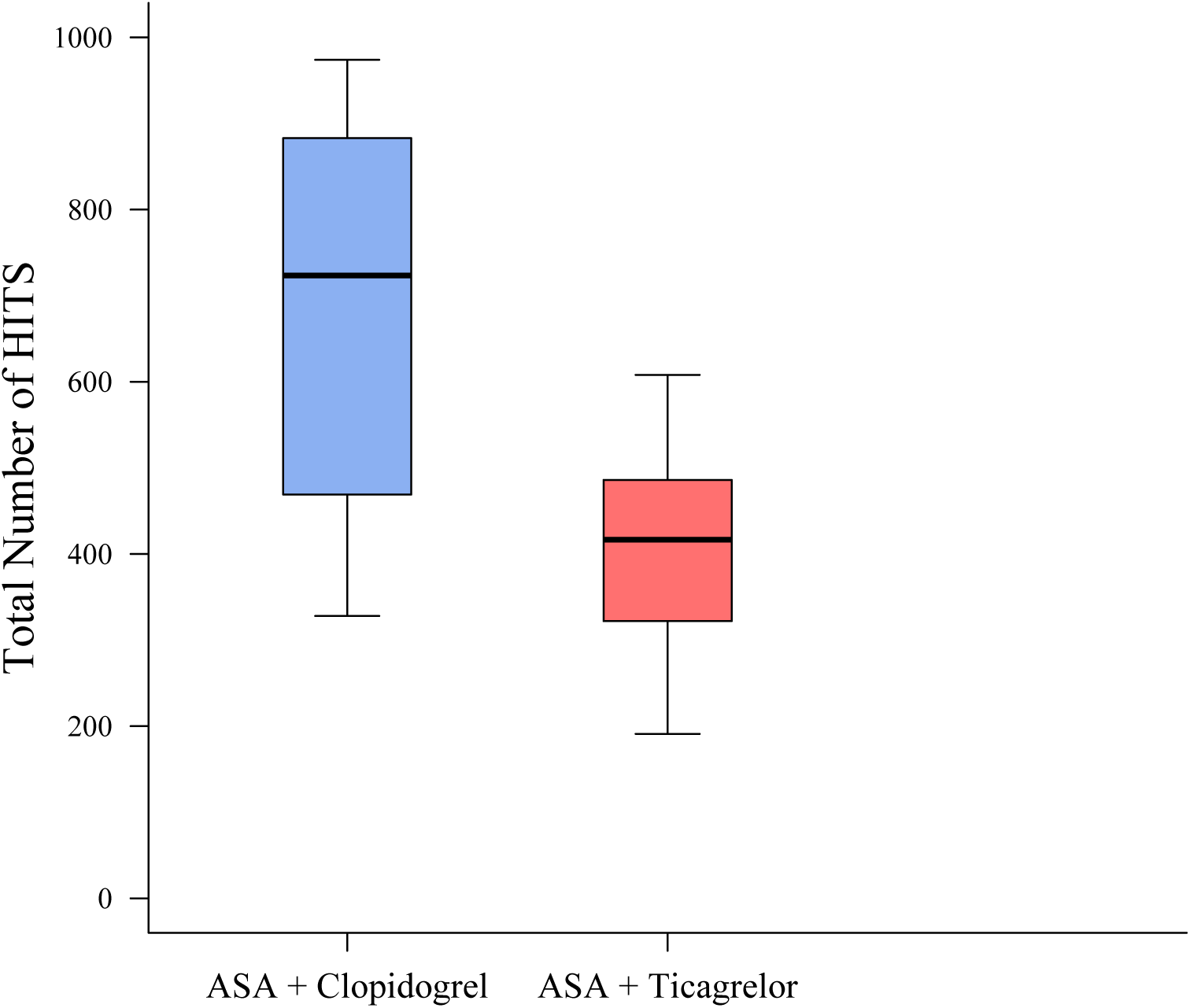
Central Illustration. Total Number of High Intensity Transient Signals in the Aspirin and Clopidogrel and Aspirin and Ticagrelor Group.

In the final regression model, patients who received aspirin and ticagrelor had, on average, 256.7 (p <0.001) fewer total procedural HITS than did those who received aspirin and clopidogrel. In addition, participants had on average two more HITS for each minute increase in the duration of the procedure **(Table 4)**.

**Table 4.**
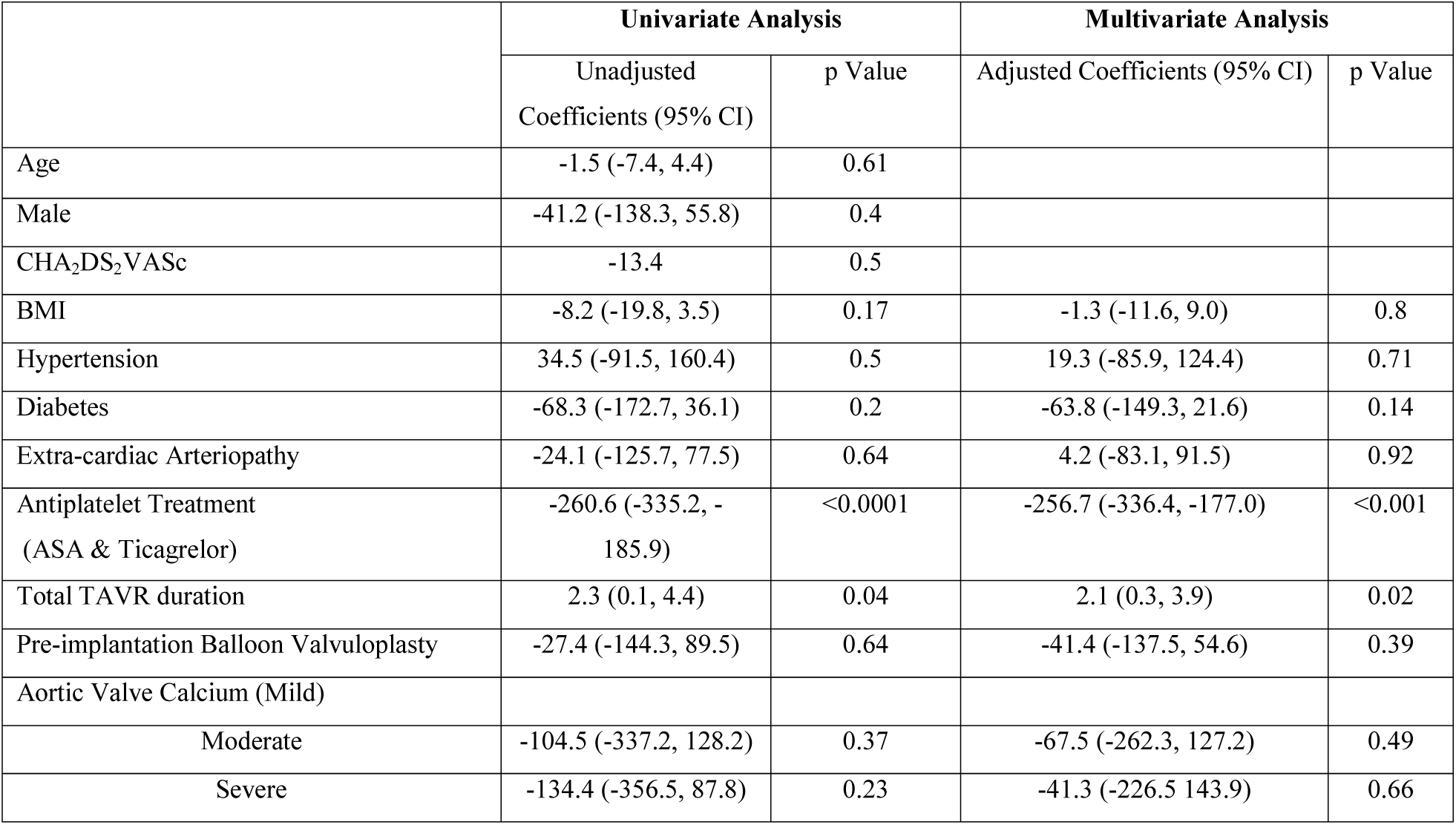
Univariate and Multivariate Regression Analysis

After adjusting for the same covariates, further analysis revealed that a one unit increase in PRU was associated with, on average, one more total HITS (p<0.001), whereas increasing platelet inhibition by one percent was associated with, on average, 3 fewer total HITS (p<0.001).

### Secondary outcomes

#### i) HITS during TAVR and at discharge

There were no HITS in either group during the 10 minutes before the procedure. The number of HITS was significantly lower in the ticagrelor group than in the clopidogrel group for all phases of the procedure. For example, during phase 1 (defined as the period from femoral vessel puncture until the transcatheter introduction of the valvuloplasty balloon) patients in the ticagrelor group had 170 [99.75, 222.50] HITS compared to 289 [161.5, 412.8] in the clopidogrel group **(Table3) (Figure 3)**.

**Figure 3.**
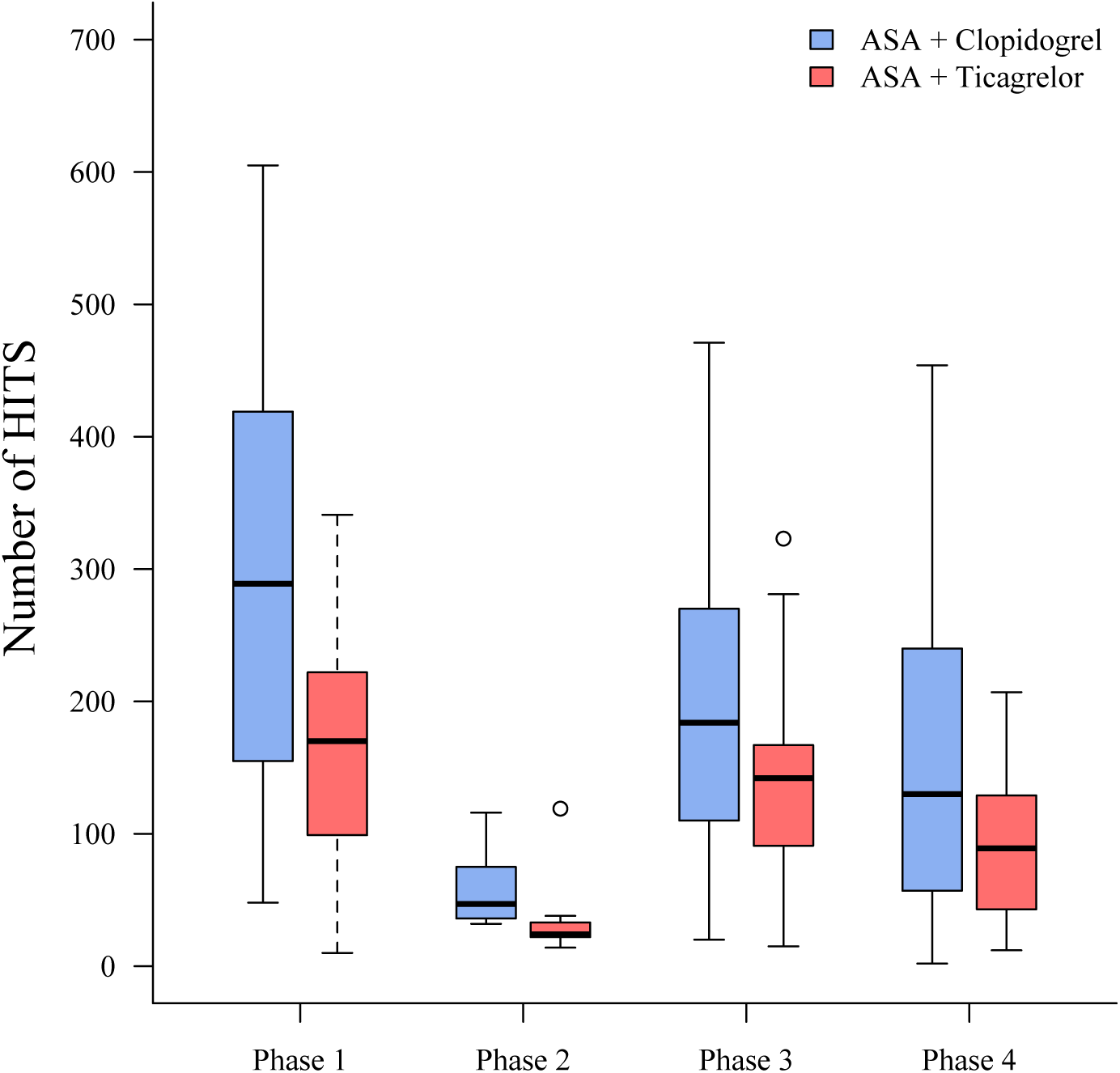
Number of HITS per Treatment Group for each Procedural Phase.

At discharge, 71 of the 84 patients underwent transcranial Doppler study. Ten patients were excluded because they had reached a safety outcome (see below) which resulted in discontinuation of the study drug, while three refused to undergo the discharge study procedures. At discharge, two participants in the clopidogrel group had six and four HITS, whereas none of the participants in the ticagrelor group had any HITS. (p=0.034).

#### ii) Platelet inhibition

Residual platelet reactivity was evaluated in 90 patients on the day of the procedure and in 74 patients at discharge. The antiplatelet regimen was discontinued in ten patients prior to discharge because they had reached a safety outcome, three patients refused to undergo the discharge study procedures, and the PRU values were not calculable by the VerifyNow assay in three patients.

P2Y12 reaction units were significantly lower on the day of the procedure in the aspirin and ticagrelor group, 26 [10, 74.5] PRU, than in the aspirin and clopidogrel group, 207.5 [120-236.2] PRU, p<0.001, with inhibition percentage of 89.0% [73.3, 96.8] and 12% [0, 30.5] in the ticagrelor and clopidogrel groups respectively (p<0.001). Linear regression showed that patients on ticagrelor had on average 130.9 fewer PRU than did those on clopidogrel. Hematocrit also significantly affected PRU, with a one-unit increase reducing the average number of HITS by four.

At discharge the median P2Y12 reaction units were also significantly lower in the aspirin and ticagrelor group, 50 [9,100] than in the aspirin and clopidogrel group, 204 [125,247], p< 0.001. The percentage of platelet inhibition was 20% [4.5, 36.7] and 81% [69, 96] in the ticagrelor and clopidogrel groups respectively, (p< 0.001).

Base PRU, measured by the extent of platelet aggregation induced by thrombin receptors prior to study drug administration was not different between the two groups: 253.5 [221, 274.8] (aspirin and clopidogrel) and 249.0 [233.8, 280] (aspirin and ticagrelor), p= 0.34.

#### iii) Extent of Platelet Inhibition and HITS

In our study, there was a significant correlation between the number of HITS and both platelet inhibition (r= 0.5, p<0.005) and percent of platelet inhibition (r= -0.5, p<0.005) **(Figure 4)**. After adjusting for diabetes, extracardiac arteriopathy, hypertension, BMI, aortic valve calcium content and pre-implantation aortic balloon valvuloplasty, a one unit increase in PRU was associated with, on average, one more procedural HITS (p<0.001), whereas increasing platelet inhibition by one percent was associated with, on average, 3 fewer total HITS (p<0.001). In both models, similarly to the main model, increasing the duration of the procedure by 1 minute was associated with, on average, two more procedural HITS.

**Figure 4.**
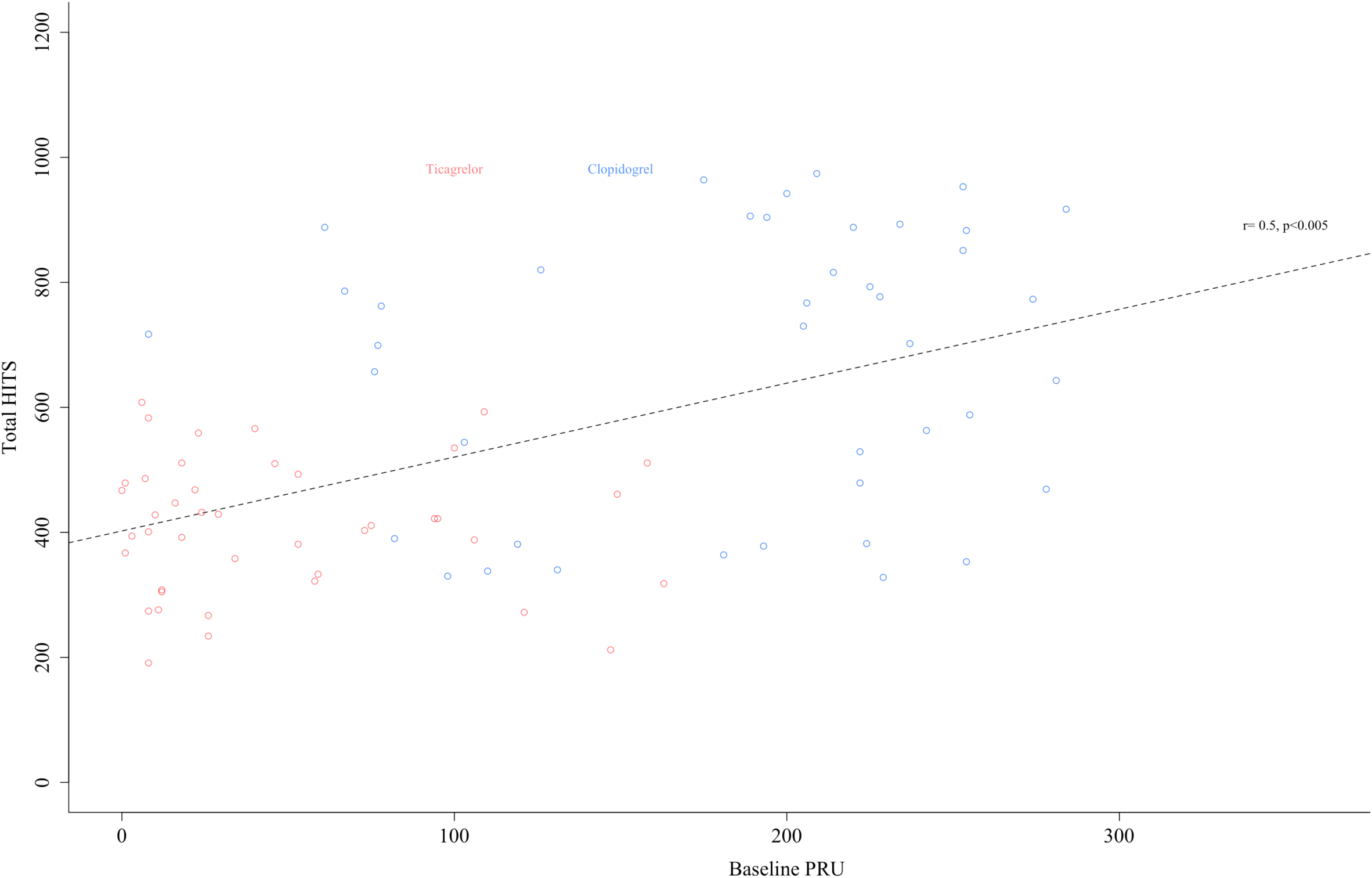
Platelet Inhibition and High Intensity Transient Signals.

### Safety Outcomes

#### Adverse Events and Serious Adverse Events during Hospitalization

No intraprocedural deaths occurred. Three SAEs, consisting of one cerebrovascular event, one pericardial effusion induced by temporary pacemaker placement and treated with pericardiocentesis, and one paraplegia attributed to injury of the Adamkiewicz artery from a descending aortic dissection, and no AEs, occurred in the clopidogrel group. Four AEs, consisting of one transient thrombocytopenia, two episodes of new onset atrial fibrillation, and one shortness of breath and four SAEs, consisting of one episode each of haematuria, haemorrhage, thrombocytopenia associated with significant anaemia requiring transfusion, and a pocket hematoma after pacemaker implantation occurred in the ticagrelor group. There were no significant differences in the number of AEs or SAEs between the two groups. (p=0.124 and 1 respectively).

#### Bleeding complications

Bleeding complications did not differ significantly between the two groups. During hospitalization, 4 patients of the ticagrelor group and 3 of the clopidogrel group developed bleeding complications according to VARC 2 criteria. In the ticagrelor group we observed two minor, one major and one life threatening bleeding, whereas in the clopidogrel group, we observed two minor and one major bleeding **(Figure 5)**.

**Figure 5.**
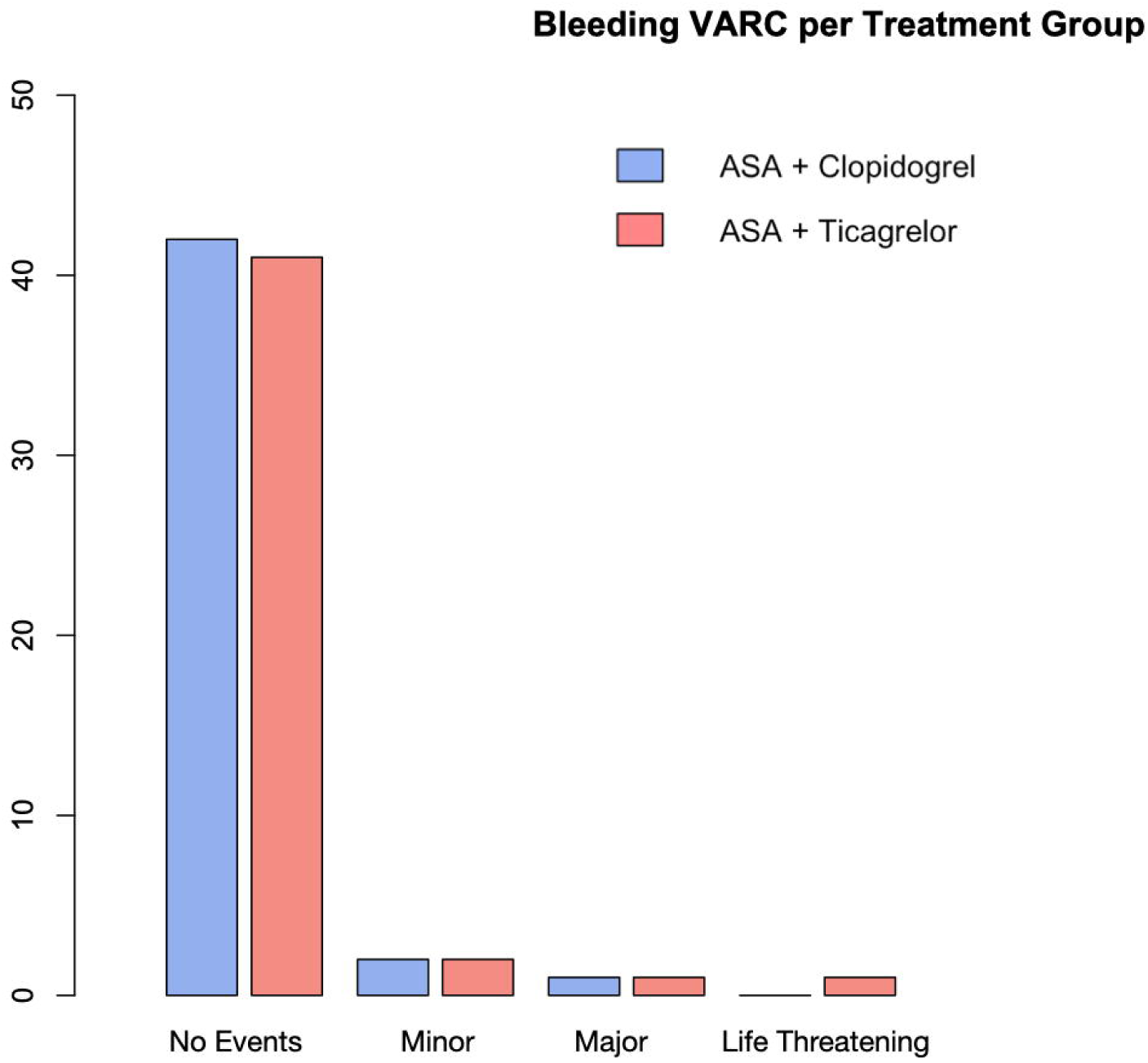
Bleeding Complications According to VARC-2 Criteria. VARC= Valve Academic Research Consortium.

#### Cerebrovascular events

Mini-Mental scores prior to the procedure were 26.78 ± 3.09 for the aspirin and ticagrelor group and 27.37±2.07 for the aspirin and clopidogrel group (p=0.317). There was one cerebrovascular event in the clopidogrel group as noted in the serious adverse events section. At hospital discharge, Mini-Mental scores were 26.54± 3.01 and 27.18±2.65 in the ticagrelor and clopidogrel groups respectively and did not differ (p=0.316).

#### Follow up at 30 and 90 days

One patient who developed, as previously mentioned, paraplegia attributed to injury of the Adamkiewicz’s artery from a descending aortic dissection, died. No cerebrovascular events were recorded at 30- and 90-day follow-up. Two patients discontinued the study medication, one for personal reasons and the other due to hematuria. Neurological assessment did not reveal any abnormalities apart from left hemiplegia in the patient who sustained the stroke. Mini-Mental Scores were not significantly different between the aspirin and ticagrelor (26.09± 3.04) and the aspirin and clopidogrel groups (27.39± 2.43, p=0.44).

### Inter-observer and intra-observer reliability

***(Supplementary Appendix)***

## DISCUSSION

To the best of our knowledge, this is the first study comparing the occurrence of cerebral microembolization, as detected by HITS in TAVR patients randomized to ticagrelor or to clopidogrel. The achieved platelet inhibition with ticagrelor was greater than with clopidogrel throughout the study. We found that the total recorded HITS during TAVR was lower in the ticagrelor than in the clopidogrel group during all phases of the procedure and that the extent of platelet inhibition was significantly correlated with the number of HITS.

Previous studies have reported a significant correlation between solid particle embolization, as detected by TCD, and aortic valve calcium score (18). Furthermore, instruments are introduced into the arterial circulation during the TAVR procedure and ascending aortic wall trauma and damage to the aortic valve tissue occur during catheter manipulation. All these are potential sites for thrombus formation augmented by activated platelets.

AHA/ACC guidelines, based on expert consensus, recommend the use of dual antiplatelet therapy for patients in the absence of a high bleeding risk. This is based on the concept that the frame of the bioprosthetic valve would behave, after its implantation, similar to that of a coronary artery stent, requiring 3 to 6 months of antiplatelet treatment while endothelization of the metallic valve frame is ongoing (19).

Of course, questions persist regarding the frame that is not in contact with the aortic wall and which remains uncovered and potentially thrombogenic. Single versus dual antiplatelet therapy has been compared previously in randomized controlled trials (7, 8, 20). The ARTE trial documented a trend towards increased frequency of life-threatening bleedings with dual antiplatelet therapy, with similar rates of MI and stroke in the two groups. Similarly, the POPular-TAVI trial documented that aspirin alone was associated with fewer bleeding complications and no difference in the risk for death, stroke and MI compared to dual antiplatelet therapy (20). However, none of these studies evaluated the degree of microembolizations to the cerebral circulation during the procedure. Nevertheless, neuroimaging studies after TAVR have documented new “silent” cerebral emboli in two of the patients, but are rarely associated with clinical events (21).

Despite the greater platelet inhibition with ticagrelor, there were no statistical differences in life threatening, disabling, major, or minor bleedings. The low incidence of bleedings observed in our study may be attributed to several factors: (1) The low HAS-BLED score of 1.75-1.88; (2) strict exclusion criteria and thorough gastrointestinal screening prior to TAVR; (3) several procedural factors including the small sheath size, CT guided arterial access, and the use of closure devices at the end of the procedure; and (4) both centers had extensive experience in TAVR procedures.

In our study we observed a high number of HITS during the TAVR procedure. We intentionally used the lowest recommended backscatter threshold to increase the sensitivity of detecting microemboli (3 dB) (12). This cut off value excludes the backscatter of the ultrasound from normal flowing blood and detects both solid and gaseous emboli.

We did not observe any significant clinical neurological differences between the two groups. However, previous studies tracing clinically silent lesions with diffuse weighted MRI, have identified that some of these lesions on follow up scans demonstrate structural tissue changes and reactive astrogliosis, thought to be a response to injury (22).

In addition, the CHA_2_DS_2_VASc score, a prognostic indicator for stroke and mortality at one year in patients undergoing TAVR, was relatively low in our study population (23). This is consistent with clinical studies that report low overt cerebrovascular events during TAVR (2%-3%). Similarly, the results of the Mini-Mental status examinations in the two groups did not differ. Finally, the fact that significant carotid artery disease was an exclusion criterion also may have contributed to the low incidence of cerebrovascular events.

### Study limitations

This study was an open label randomized clinical trial and therefore patients and the physicians performing the procedure were aware of the assigned antiplatelet regimen. However, those who were measuring and interpreting TCD data and those performing the neurological assessments were blinded to the antiplatelet regimen. Another limitation of our study is the lack of platelet function assessment prior to the administration of ticagrelor or clopidogrel. However, the initial platelet function assessment in our trial included measurement of base platelet reactivity, which assesses the rate and extent of platelet aggregation from the thrombin receptors, specifically the PAR-1 and PAR-4 receptors (base-PRU), a measure which is independent of P2Y12 inhibition and did not differ between the two groups. In addition, the randomization process limits the likelihood of possible non-recognized potential confounders within the two study populations.

The location of microemboli is also unknown due to the lack of imaging modalities in our study and might have involved non-eloquent areas of the brain. Therefore, we could not document the degree of injury that was produced in each group and correlate the differences in total HITS observed by the transcranial Doppler studies to actual cerebral lesions. Since a known limitation of the TCD method is that it is operator dependent, the same operator performed all the examinations of the study for each patient. In addition, the sample size of our study and the duration of follow-up were not designed to assess any differences in clinical events between the two groups.

## CONCLUSIONS

In patients undergoing TAVR, with characteristics similar to those enrolled in our study, the use of ticagrelor instead of clopidogrel, in addition to low dose aspirin, is an effective way to reduce microemboli in the cerebral circulation. The use of ticagrelor instead of clopidogrel was not associated with an increased risk of bleeding or adverse events peri-procedurally, or during the follow up period. Further studies are required to study this approach in other patient populations.

## CLINICAL PERSPECTIVES

What is known?

- Cerebral microembolic events are common in patients undergoing TAVR and can be detected using transcranial Doppler imaging.

What is new?

- The incidence of cerebral microemboli, as detected by TCD during TAVR, was significantly correlated with the extent of platelet inhibition and was lower in patients randomized to ticagrelor and aspirin than in those randomized to clopidogrel and aspirin, without increasing the risk of bleeding complications.

What is next?

- Further trials are required to evaluate other antiplatelet regimens during and after TAVR and define the risk benefit balance between cerebrovascular events and bleeding complications, especially within groups of different surgical and clinical characteristics.

## Data Availability

The data and analytical methods that support the findings of this study are available from the corresponding author on reasonable request.

## Acknowledgements

We would like to sincerely thank Dr. Gary Gerstenblith for his careful review of the manuscript and Dimitra Latsou, PhD from Pharmecons Easy Access for her contribution regarding data analysis.

## LIST OF ABBREVIATIONS

ACS: Acute Coronary Syndrome
ASA: Acetylsalicylic Acid
MRI: Magnetic Resonance Imaging
PRU: P2Y12 reaction units
TAVR: Transcatheter Aortic Valve Replacement
TCD: Transcranial Doppler
SAVR: Surgical Aortic Valve Replacement
VARC: Valve Academic Research Consortium

## Notes

### Competing Interest Statement

The authors have declared no competing interest.

### Clinical Trial

ClinicalTrials.gov Identifier: NCT02989558

### Funding Statement

Funding/ Support: The study was supported by Astra Zeneca.
Role of the Funder/Sponsor: Astra Zeneca, the funder of the study, had no role in the collection, management, or interpretation of the data, or the statistical analysis; the funder reviewed the manuscript but was not involved in the writing or approval of the manuscript or the decision to submit the manuscript for publication.
Dr. Manolis Vavuranakis is a proctor for Medtronic and Abbott Laboratories. Dr. Toutouzas is a proctor for Medtronic and has received research grants from Medtronic, Bayern and Pfizer. Dr. Voudris is on the Medtronic Advisory Board. The remaining authors have no disclosures.

### Author Declarations

The study was conducted in accordance with the ethical principles for medical research of the Declaration of Helsinki, International Conference on Harmonization (ICH) /Good Clinical Practice (GCP), the European Union Clinical Trials Directive, and Greek legislation. The study protocol was approved by the hospitals Ethics Committee and Institutional Review Board, the National Ethics Committee (NEC), and the National Organization of Medicines (NOM). Safety updates were provided according to local requirements, including SUSARs (Suspected Unexpected Serious Adverse Reactions).

