## Supplementary Appendix for "Ticagrelor vs Clopidogrel: the Impact of Platelet Inhibition on Cerebrovascular Microembolic Events during TAVR"

**SUPPLEMENTARY APPENDICES**

### Table of Contents

Appendix A- Inclusion and Exclusion Criteria

Appendix B- Flowchart of Study Procedures

Appendix C- Statistical analysis, sample size and power calculations

Appendix D- Inter-observer and intra-observer reliability

Appendix E- Procedural Stages

Appendix F- Adverse Events (AEs) and Serious Adverse Events (SAEs) definitions

Supplementary Appendix References

**Appendix A- Inclusion and Exclusion Criteria**

### 1.1 Inclusion Criteria

Subjects eligible for study inclusion met all of the following criteria:

1 The subject (or where unable to do so, their legally acceptable representative) must be able to provide written informed consent prior to any study specific criteria, stating that he or she understands the purpose of and the procedures required for the study and is willing to participate in the study.
2 Female and/or male subjects aged 18 years or older.
3 High risk (EuroSCORE^[1]^ ≥18 or considered inoperable) for surgical aortic valve replacement.
4 Is expected to benefit from the placement of TAVI.
5 Does not suffer from any disease or condition that would limit his/her life expectancy of <6 months.
6 The subject must be willing and able to adhere to the prohibitions and restrictions specified in this protocol.
7 Subjects must meet the following laboratory results from Visit 0 for study inclusion^[2]^:

a. Hemoglobin ≥ 10 g/dL

b. Platelets ≥ 100 X 10^3^ cells/μL

c. Absolute neutrophil count (ANC) ≥ 1000 cells/μL

d. Aspartate aminotransferase (AST), alanine aminotransferase (ALT), and alkaline phosphatase (ALP) levels must be within 1.5 times the upper limit of normal (ULN) range for the laboratory conducting the test.

e. Total bilirubin ≤ 2x ULN.

### 1.2 Exclusion Criteria

Subjects will not enter the study if any of the following exclusion criteria are fulfilled:

1. Have scheduled any elective surgery in the next 4 months following screening procedures.
2. History of hypocoagulopathies.
3. Previous thromboembolism or known hypercoagulopathy (acquired or congenital).
4. Antiplatelet therapy, other than ASA, within 7 days before randomization that cannot be discontinued due to the underlying disease.
5. History of gastric or duodenal ulcer disease verified by endoscopy or barium meal double-contrast technique within 3 months prior to randomization.
6. Moderate or severe hepatic impairment.
7. Known hypersensitivity to any of the investigational products or their components.
8. Presence of any other clinically significant disease or disorder which, in the opinion of the Investigator (by its nature or by being inadequately controlled), might put the subject at risk due to participation in the study, or may influence the conduct or the interpretation of the results of the study or the subject’s ability to complete the study.
9. Any identified contraindication for using ticagrelor, clopidogrel or aspirin.
10. History or recent findings of atrial fibrillation.
11. Significant carotid artery disease on either internal carotid artery, as defined by a >50 % diameter reduction on carotid ultrasonography.
12. Unwillingness to receive or intolerant to any blood products.
13. Previous trauma or surgery to either femoral artery.
14. Major surgical procedure or trauma within the 30 days prior to enrolment.
15. Mechanical heart valve (any location).
16. Mitral or aortic bioprosthetic valve.
17. The subject is an employee of the investigator or study site, with direct involvement in the proposed study or other studies under the direction of that investigator or study site, as well as family members of the employees or the investigator.
18. Reproductive status:
    a. Women who are pregnant or planning a pregnancy within 1 month of the end of study;
    b. Women who are breastfeeding;
    c. Women of childbearing potential who are unwilling or unable to use two highly effective methods of birth control to avoid pregnancy for the entire study period, as evaluated by the Investigator. Women who are not of childbearing potential are those that have a history of hysterectomy, bilateral oophorectomy, or are postmenopausal with no history of menstrual flow for ≥ 12 months prior to the screening Visit.

Highly effective methods of birth control are defined as:

- Hormonal: established use of oral, implantable, injectable, or transdermal hormonal methods of conception;
- Placement of an intrauterine device;
- Placement of an intrauterine system;
- Mechanical/barrier method of contraception: condom or occlusive cap (diaphragm or cervical/vault cap) in conjunction with spermicide (foam, gel, film, cream or suppository);
- When used alone, the diaphragm and condom are not highly effective forms of contraception. Therefore the use of additional spermicides provides more effective birth control. However, spermicides alone may not be effective in preventing pregnancy and must be used with a barrier such as a condom or diaphragm.
- Surgical sterilization of male partner (with the appropriate post-vasectomy documentation of the absence of sperm in the ejaculate; for female patients on the study, the vasectomized male partner should be the sole partner for that patient) in conjunction with spermicide, condom, or diaphragm;
- Sexual true abstinence: when this is in line with the preferred and usual lifestyle of the subject. Periodic abstinence (e.g., calendar, ovulation, symptothermal, post-ovulation methods) and withdrawal are not acceptable methods of contraception.

19. International normalized ratio (INR) ≥ 2 on the day before the TAVI procedure.
20. History of hemorrhagic stroke, intracranial, intraocular, spinal, retroperitoneal or atraumatic intra-articular hemorrhage, intracerebral mass or aneurysm, or arteriovenous malformation.
21. Severe left ventricular dysfunction (left ventricular ejection fraction<15%).
22. Severe aortic regurgitation or mitral regurgitation, according to the 2012 European Society of Cardiology and the European Association for Cardio-Thoracic Surgery guidelines (See Appendix 2)
23. Hemodynamic instability (e.g. requiring inotropic or intra-aortic balloon pump support) within 2 hours of the procedure.
24. Subject is dialysis dependent.
25. Any condition requiring the use of anticoagulants that cannot be stopped for the duration of the study.
26. Acute myocardial infarction, major surgery or any therapeutic cardiac procedure (other than balloon aortic valvuloplasty) within 30 days.
27. Gastrointestinal or genitourinary bleed within 30 days.
28. Absolute contraindications or allergy to iodinated contrast that cannot be pre-medicated.
29. Treatment with other investigational drugs (including investigational vaccines) or devices within the 30 days preceding enrolment or planned use of other investigational drugs or devices before the end of the study.
30. Known alcohol or drug abuser.
31. The subject has received or scheduled to receive for the study period strong CYP3A inhibitors (ketoconazole, itraconazole, voriconazole, telithromycin, clarithromycin, erythromycin, nefazadone, ritonavir, saquinavir, nelfinavir, indinavir, atanazavir, over 1 litre daily of grapefruit juice), CYP3A substrates with a narrow therapeutic index (cyclosporine, quinidine, simvastatin at doses >40 mg daily, lovastatin at doses >40 mg daily), and strong CYP3A inducers (rifampin/rifampicin, phenytoin, carbamazepine, phenobarbital).

**Appendix B- Flowchart of Study Procedures**

|  | **Procedures** | **Screening Period** | **Baseline** | **In-hospital period** | | **End Of Treatment & End of Study** | **Follow up** |
| --- | --- | --- | --- | --- | --- | --- | --- |
| **Time (days)** |  | **-60 to -11** | -10 to -1 | **0 (TAVI)** | 4-7 (discharge) | **90** | 120 |
| **Eligibility** | Informed Consent | **X** |  |  |  |  |  |
|  | Inclusion/Exclusion Criteria | **X** | X |  |  |  |  |
|  | CT scan | **X** |  |  |  |  |  |
|  | Transthoracic echocardiography | **X** |  |  |  |  |  |
|  | Ultrasound of the carotid arteries | **X** |  |  |  |  |  |
|  | EuroSCORE assessment | **X** |  |  |  |  |  |
|  | Medical/Surgical history | **X** | X |  |  |  |  |
|  | Demographics |  | X |  |  |  |  |
| **Safety** | Concomitant medication review |  | X | **X** | X | **X** |  |
|  | Adverse Events collection |  | X | **X** | X | **X** | X |
|  | Bleeding events recording (VARC-2) |  | X | **X** | X | **X** | X |
|  | Complete Physical Examination |  | X |  |  | **X** |  |
|  | Limited Physical Examination |  |  | **X** | X |  |  |
|  | Vital signs |  | X | **X** | X | **X** |  |
|  | 12-Lead ECG | **X** | X | **X** | X |  |  |
|  | Hematology & biochemistry |  | X |  | X |  |  |
|  | Pregnancy test (urine or serum) |  | X |  |  |  |  |
| **Study procedures** | Randomization |  | X |  |  |  |  |
|  | Neurologic assessment |  | X |  | X | **X** |  |
|  | Transcranial Doppler |  |  | **X** | X |  |  |
|  | Blood sample for induced platelet inhibition^§^ assesment |  |  | **X** | X |  |  |
|  | Investigational products dispensed |  | X |  |  |  |  |
|  | Investigational products collected |  |  |  |  | **X** |  |

**Appendix C- Statistical analysis, sample size and power calculations**

Size of study population was calculated as described by Pocock SJ (1) utilizing the formula

$n=\frac{2\sigma^{2}}{{(\mu_{2}-\mu_{1})}^{2}}\cdot f(\alpha,\beta)$, where f(α,β) is a constant given from a reference table (for type I error (α) = 0.05 and type II error (β) = 0.10, f(α, β) = 10.5), $\mu_{2}=$ mean value of HITS during TAVR in reference studies for TAVR subjects under clopidogrel plus ASA, $\mu_{1}=$ the targeted mean value of HITS during TAVR for subjects treated with ticagrelor plus ASA.

In particular, we use previously presented data (2) showing that the mean total procedural HITS during transfemoral study were 658 ±275. Therefore: $\mu_{1}=$658 and σ=275 were utilized in the formula.

Since the average % platelet inhibition is reported to be about 30% higher with ticagrelor (89.9%) than it is for clopidogrel (67.4%) (3) we tested the hypothesis that the ticagrelor regimen would achieve a 30% reduction in total HITS when compared with the clopidogrel regimen. The needed sample size for each group was calculated to be 40 patients. In order to adjust for possible poor-quality Doppler and under the assumption of ~10% drop out rate, 45 subjects were finally recruited in each group, leading to a total of 90 participants.

**Appendix D- Inter-observer and intra-observer reliability**

Inter-observer reliability: Total number of HITS of 10 randomly selected patients were measured by the two independent observers. The Bland-Altman method was used to depict inter-observer difference. The mean difference of measurement between two observers was -5.7. The correlation coefficient between two independent observers was r=0.98, p<0.001.

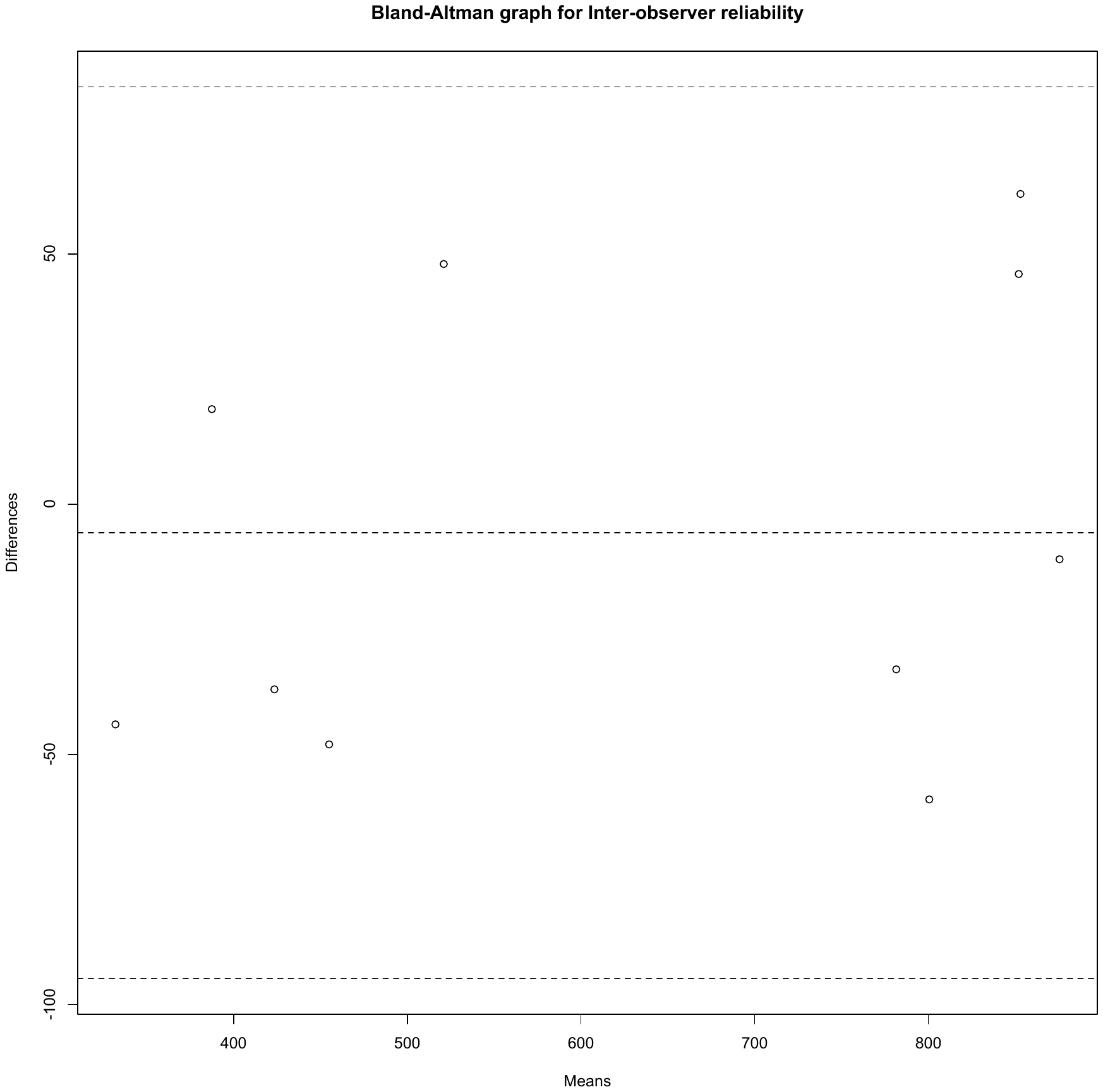

Intra-observer reliability: The first observer measured the total number of HITS of eight randomly selected patients twice. The mean difference between the measurements was 3 and the correlation coefficient was r=0.991, p<0.001.

Similarly, the second observer also measured the total number of HITS of eight randomly selected patients twice. The mean difference of measurement was 0,75 and the correlation coefficient was r=1.000, p<0.001.

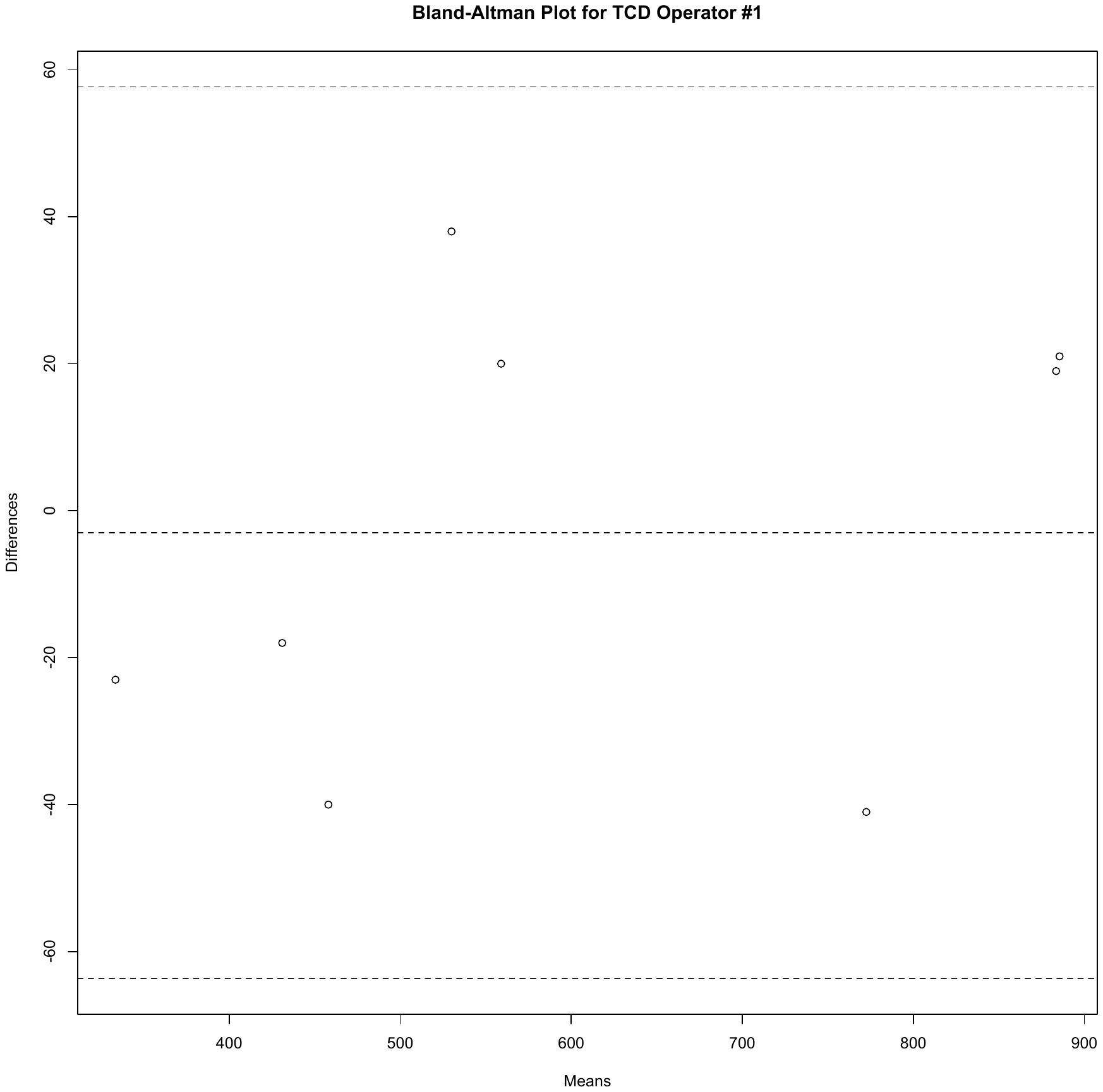

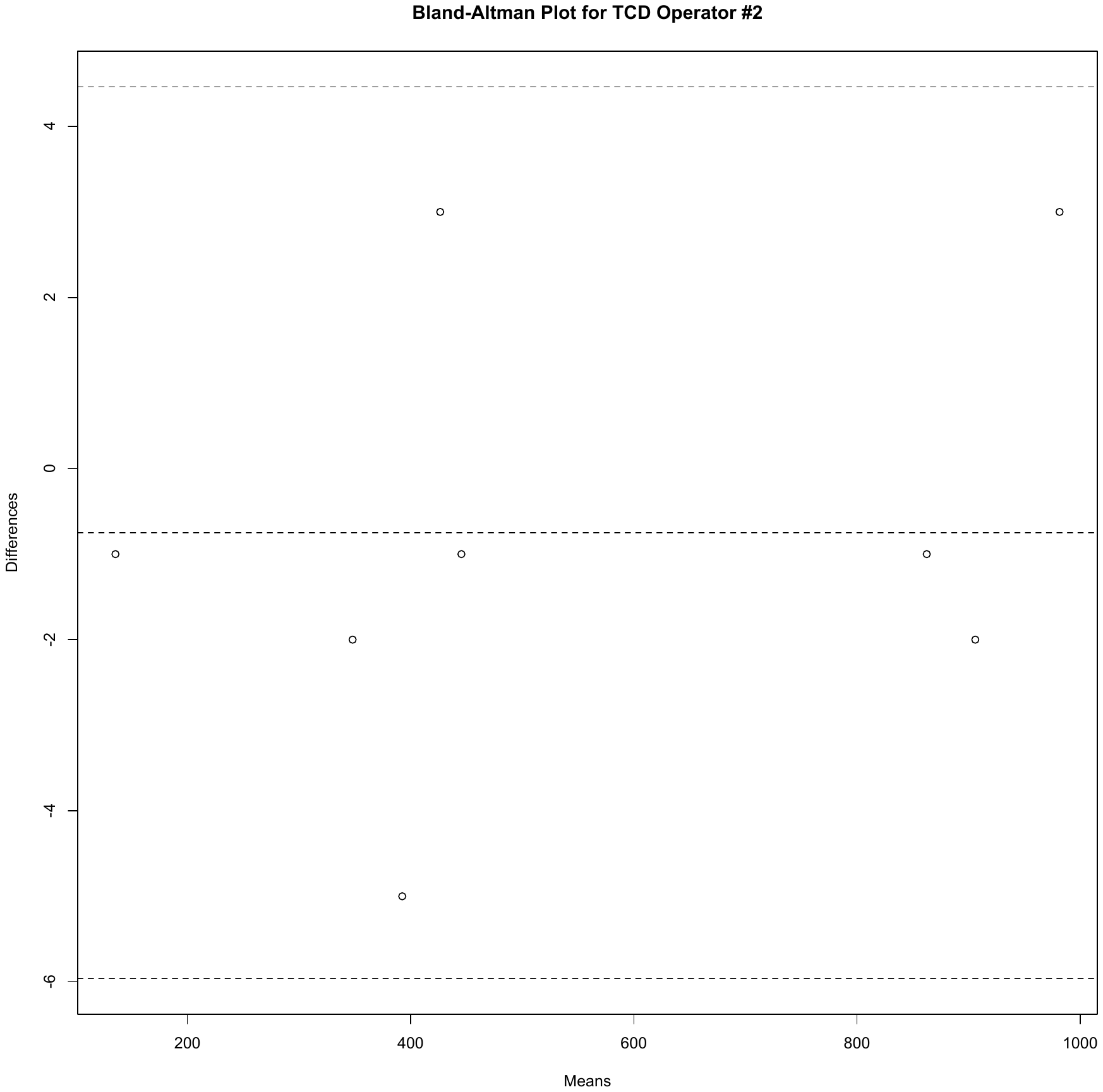

**Appendix E- Stages of the procedure**

*Phase 1*: defined as the period from first femoral vessel puncture until the transcatheter introduction of the valvuloplasty balloon;

*Phase 2*: defined as the period from the transcatheter introduction of the valvuloplasty balloon until the transcatheter introduction of the self-expandable valve;

*Phase 3*: prosthesis deployment, defined as the period from the transcatheter introduction of the self-expandable valve until the removal of delivery system;

*Phase 4*: defined as the period from the removal of the valve delivery system until the closure of the access site, also including any post-dilatation episodes.

**Appendix F- Adverse Events (AEs) and Serious Adverse Events (SAEs) definitions**

#### Definition of Adverse Events

An adverse event is the development of an undesirable medical condition or the deterioration of a pre-existing medical condition following or during exposure to a pharmaceutical product, whether or not considered causally related to the product. An undesirable medical condition can be symptoms (e.g. nausea, chest pain), signs (e.g. tachycardia, enlarged liver) or the abnormal results of an investigation (e.g. laboratory findings, electrocardiogram). In clinical studies, an AE can include an undesirable medical condition occurring at any time, including run-in or washout periods, even if no study treatment has been administered.

#### Definitions of Serious Adverse Events

A serious adverse event is an AE occurring during any study phase (i.e. run-in, treatment, washout, follow-up), that fulfils one or more of the following criteria:

- Results in death
- Is immediately life-threatening
- Requires in-patient hospitalization or prolongation of existing hospitalization
- Results in persistent or significant disability/incapacity or substantial disruption of the ability to conduct normal life functions
- Is a congenital abnormality or birth defect
- Is an important medical event that may jeopardize the subject or may require medical intervention to prevent one of the outcomes listed above.

1. Visit [www.euroscore.org/calculators](http://www.euroscore.org/calculators) for a downloadable or online tool for calculation of this assessment.
2. A one-time repeat is allowed and the Investigator may consider the subject eligible if the previously abnormal laboratory test result is within acceptable range on repeat testing.
